# Cardiovascular risk management in the elderly with type 2 diabetes prior to death: A Danish nationwide register study of discontinuation patterns

**DOI:** 10.1101/2024.02.23.24303300

**Authors:** Vanja Kosjerina, Stine H. Scheuer, Bendix Carstensen, Marit E. Jørgensen, Birgitte Brock, Hanne R Christensen, Gregers S Andersen, Jørgen Rungby

## Abstract

**Background:** Cardioprotective medication usage among elderly individuals with type 2 diabetes (T2D) is prevalent, however, the degree and timing of discontinuation is unknown. This study aims to describe the extent, timing, and secular changes of discontinuation of cardioprotective medication in elderly with T2D.

**Methods:** In this register-based cohort study all individuals with T2D, deceased between 2006-2018 at an age of 80 years or older, were identified through Danish nation-wide registers. We followed the population backward in time, from death to last intake of antihypertensive, lipid-lowering, and antithrombotic medication. Poisson regression models were used to model rates of discontinuation prior to death while binomial models were used to estimate the proportion on medication at time of death.

**Results:** We identified 52,523 individuals (55% women) with a mean (SD) age at T2D diagnosis of 77.3 (7.8) years and median (Q1-Q3) age at death of 86.5 [83.3-90.3] years. The proportion on any antihypertensive and antithrombotic medication was high (approximately 78% [95% CI 77-78] and 48% [95% CI 47-49%] at time of death, respectively) and increased slightly with increasing calendar year of death. Discontinuations occurred predominantly in the last year of life but were initiated earlier for individuals who died in recent calendar years. However, for angiotensin-converting enzyme, thiazides, and acetylsalicylic acid we found a more continuous discontinuation in the decade prior to death in the recent calendar years of death. The proportion on statins increased markedly with calendar year of death, peaking at 34% [95% CI 33-35%] in 2016 with a subsequent decrease to 29% [95% CI 28-30%] in 2018. Discontinuation patterns shifted from predominantly occurring in the last years of life to continues discontinuation in the decade before death.

**Conclusion:** Our results suggest an intensification with cardioprotective medication among elderly with type 2 diabetes and a more pronounced discontinuation during the last years of life. This may be a result of increased focus on individualized-treatment-regimens.

## Introduction

One of the greatest achievements of the 21 century is a generally increased life expectancy. ^1^ Following advancements in treatment of type 2 diabetes (T2D) and the resulting decrease in excess mortality, there has been a rise in the prevalence of individuals aged 65 and above with T2D.^2^ As both age and diabetes duration are independently predictive of morbidity, the increase in elderly with T2D has led to a rise in individuals affected by multimorbidity, with cardiovascular disease (CVD) being one of the most prevalent.^3,4^ Treatment guidelines for elderly with T2D acknowledge that although glycemic management is of importance, modifying CVD risk factors is more likely to reduce both morbidity and mortality.^5^ Hence, a large proportion of the population will receive cardioprotective medication as either primary or secondary prevention.

Treatment of elderly and very elderly (>80 years) with T2D is complicated by their heterogeneity in terms of the clinical, functional, and cognitive status and the lack of robust evidence to guide treatment decisions. Guidelines are often based on extrapolated evidence from randomized control trials (RCT) where the elderly population is either underrepresented or unevenly, typically skewed toward by the healthiest subgroup.^6^ Furthermore, the high burden of multimorbidity in this population adds to the complexity of treatment and the risk for polypharmacy in the presence of multiple disease specific guidelines. Polypharmacy, with a 64% prevalence in the elderly with T2D, is associated with unfavorable outcomes such as non-adherence, risk of hospitalizations, drug-drug interaction, drug-disease interaction, adverse drug reactions and inappropriate prescriptions.^7,8^

There has been an increased focus on individualized treatment goals with medication reviews and discontinuation of medications as tools to minimize polypharmacy, improve health outcomes, and reduce untimely discontinuation (non-adherence).^9,10^ It is advised that a variety of factors should be considered when prescribing and discontinuing medication; clinical status, risk-benefit of medication, life expectancy and the individual wishes of the patient. However, the same factors have also been documented as barriers for discontinuation, with emphasis on the lack of evidence and in some cases contradictory evidence.^11^ A gap in evidence that poses a particular barrier for health care professionals is the constant relative hazard assumption implied in guidelines regarding the long-term use of cardioprotective medication, in combination with the absences of clear guidance of the timing of discontinuation.^6^ Furthermore, studies (both RCT’s and observational studies) on discontinuation have unanimously shown a reduction in potentially inappropriate medications but have failed to provide clear evidence on long-term benefits of quality of life, hospitalizations, morbidity, and mortality.^9,12^

In this interplay of increased awareness of the importance of reducing CVD risk-factors and appropriate treatment regimes in the elderly, there is little knowledge of the rates of discontinuation at various stage prior to death. Limited life expectancy can be a facilitator of discontinuation as treatment goals shift from disease control and prevention to symptom relief and comfort.^5,13^ Determining life expectancy is difficult, nevertheless, the proximity to death is of importance in the decision-making process of discontinuation. Previous studies on discontinuation rates have been conducted in selected populations in terms of their clinical status (often only overtreated), the proximity to death (either solely near end-of-life or with no mention of life expectancy), the care settings (community dwelling vs end-of-life facilities) and prior adherence to medication. Few studies have focused on the T2D population, and the follow-up times have been limited.^9,14^ Given the limited evidence on this topic we sought out to describe the extent, timing, and secular changes in discontinuation of cardioprotective medication during decade prior to death, in an elderly population with T2D deceased between 2006-2018. Furthermore, we aimed to explore how various demographical, clinical and socioeconomical factors are associated with these patterns.

## Materials and methods

### Study design and participants

In this register-based cohort study all elderly individuals in Denmark, 80 years or older at time of death, with T2D and a date of death between 01.01.2006 and 31.12.2018 were identified through cross-linkage of several nationwide administrative health registers and databases.

The Danish national healthcare registers allow for large-scale epidemiological studies with a minimum loss to follow-up and the unique personal identification number allows for cross-linkage of data on an individual level.^15^ The Danish diabetes register (DMreg) was used to identify all individuals with a T2D diagnosis. DMreg consists of information from five Danish nationwide registers and databases, and with an algorithm classifies individuals as having either type 1 or type 2 diabetes.^2^ Prescription data was retrieved from the National Prescription Registry (NPR), and contains all redeemed prescriptions in Denmark since 1995.^16^ Data on comorbidities was retrieved from the Danish National Patient Register (DNPR), the Register of Laboratory Results for Research, and the Danish Adult Diabetes Database (DADD).^17–19^ Information on demographics (income, sex, age, marital status, migrations status and level of education) was retrieved from the Populatiońs Education Register, the Danish Civil Registration System, and the Danish register of personal income and transfer payments. ^20–22^ For a detailed description of all registers used, including validity of the registers, see the supplementary material.

### Definition of outcome of interest

Cardioprotective medication included three main categories: antihypertensive, lipid-lowering, and antithrombotic medication, comprising several medication classes within each category. Date of discontinuation was defined as the date of last intake of medication. The last redeemed prescription of each medication class was extracted, and date of last medication intake was calculated as the date of last acquisition plus amount redeemed (package size multipede by number of packages) divided by the prescribed daily dose (PDD). If the PDD was not recorded, then the defined daily dose (as defined by the World Health Organization) was used as a proxy.^23^ Bothe single-substance and combination drugs were included, and combination drugs would thus contribute to more than one medication class. Finally, a variable with the date of last intake of any antihypertensive or any antithrombotic medication was constructed. For the full list of Anatomical Therapeutic Chemical codes used to classify medication classes and information of the proportion of missing PDD, see Table S1.

### Definition of covariates

Diabetes related complications were categorized as: hypertensive disease, atrial fibrillation, heart failure, ischemic heart disease, macrovascular atherosclerotic disease, cerebrovascular disease, neuropathy, diabetic kidney disease, retinopathy, and amputations. International classification of disease (ICD-10) codes, procedure codes and Nomenclature for Properties and Units codes were used to define these diseases entities and are listed in Table S2. Level of education was defined as the highest education achieved, categorized as low (primary and lower secondary), medium (upper secondary) and high (short cycle tertiary, bachelors- or master’s degree and doctoral), according to the International Standard Classification of Education (ISCED 2011). Individuals were defined as immigrants if they were born in another country than Denmark and if their parents were without Danish citizenship and born outside of Denmark. Immigrants were categorized as originating from western or non-western countries, determined by the country of origin. Income quintiles (with the first being the lowest) were calculated yearly and based on the entire Danish population.^22^ Marital status was categorized as; married, widower, divorced and unmarried. All sociodemographic covariates, except country of origin, were assessed yearly.

### Statistical Analysis

To investigate timing of discontinuation relative to time of death we did a follow-back time from death to date of discontinuation of cardioprotective medication, in the population not on medication at time of death. We censored follow-back time at 10 years before death or date of T2D diagnosis, whichever came first. The follow-back time of each individual was split in 6-month intervals along the time before death. Rates of discontinuation were modeled separately for each medication class using Poisson-likelihood. Using a binomial model, we modeled the proportion of individuals on a given medication at time of death, separately for each medication class. The effect of calendar time of death was included in both models as a smooth parametric function. The cumulative risk of medication i.e., of taking medication for the last time (as derived from the discontinuation rates) and the proportion on medication at time of death were combined to compute the proportion of the population on each medication class at a given time before and at death. The results were subdivided by calendar year of death to illustrate secular changes, presented in Figure 1. From the binomial model, the proportion on each medication class, subdivided by calendar year of death, is shown in Figure 2.

**Figure 1).**
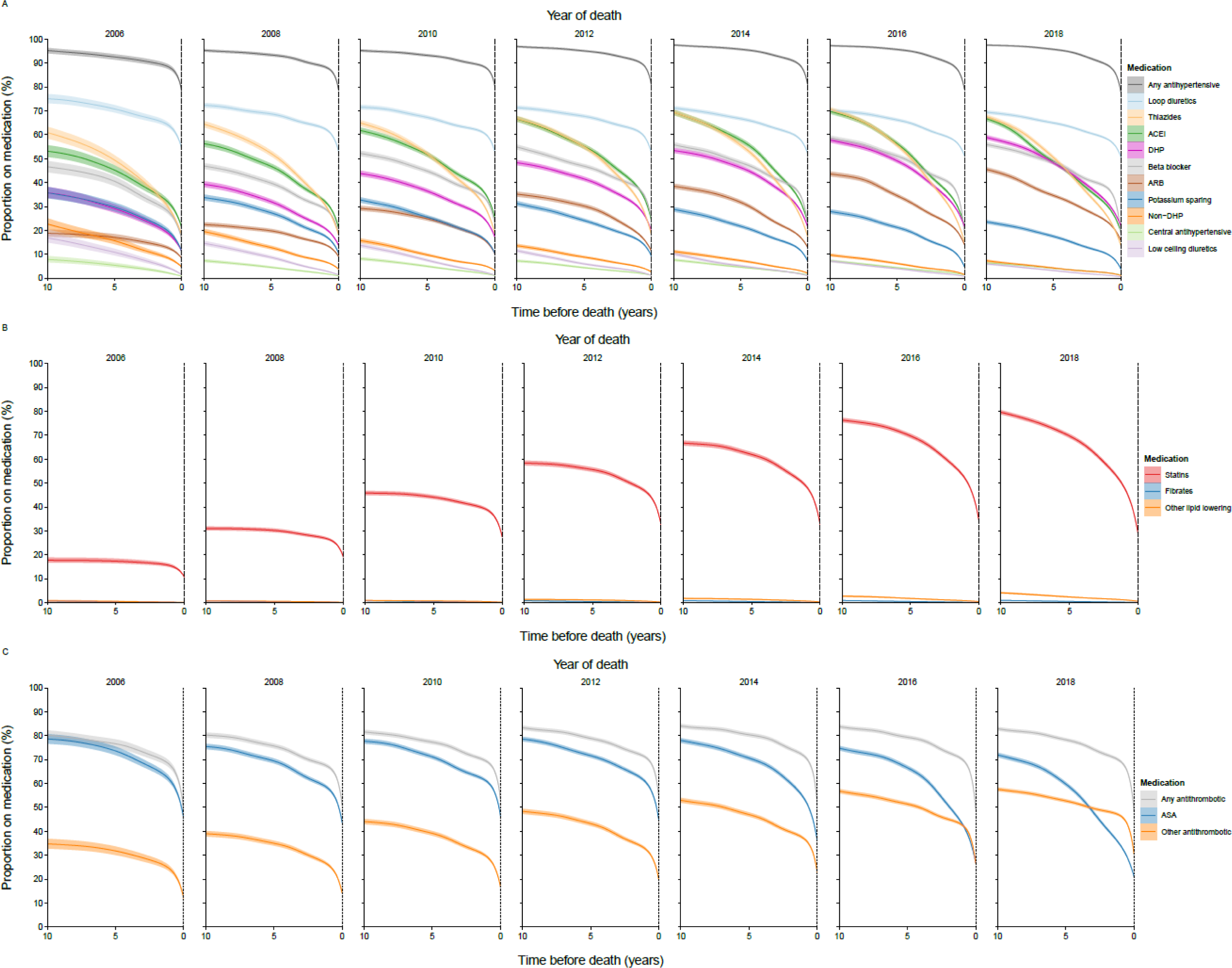
Proportion of population on A) antihypertensive, B) lipid lowering, and c) antithrombotic medication as a function of time before death, subdivided by year of death and medication class (only every other year of deaths is shown) The end point of each curve is the proportion of individuals on each type of antihypertensive/lipid-lowering/antithrombotic medication at death. Shaded areas represent 95% CI. A crude model, without additional adjustments for covariates, was used to construct the figure. Antihypertensive medication includes loop diuretics, thiazides, angiotensin-converting enzyme inhibitors (ACEi), dihydropyridines (DHP), beta blocker, angiotensin receptor blockers (ARB), potassium-sparing diuretics, non-dihydropyridines (non-DHP), central antihypertensive, and low ceiling diuretics. Lipid lowering medication includes statins, fibrates, and other lipid lowering medication. Antithrombotic medication includes acetylsalicylic acid (ASA) and other antithrombotic medication.

**Figure 2).**
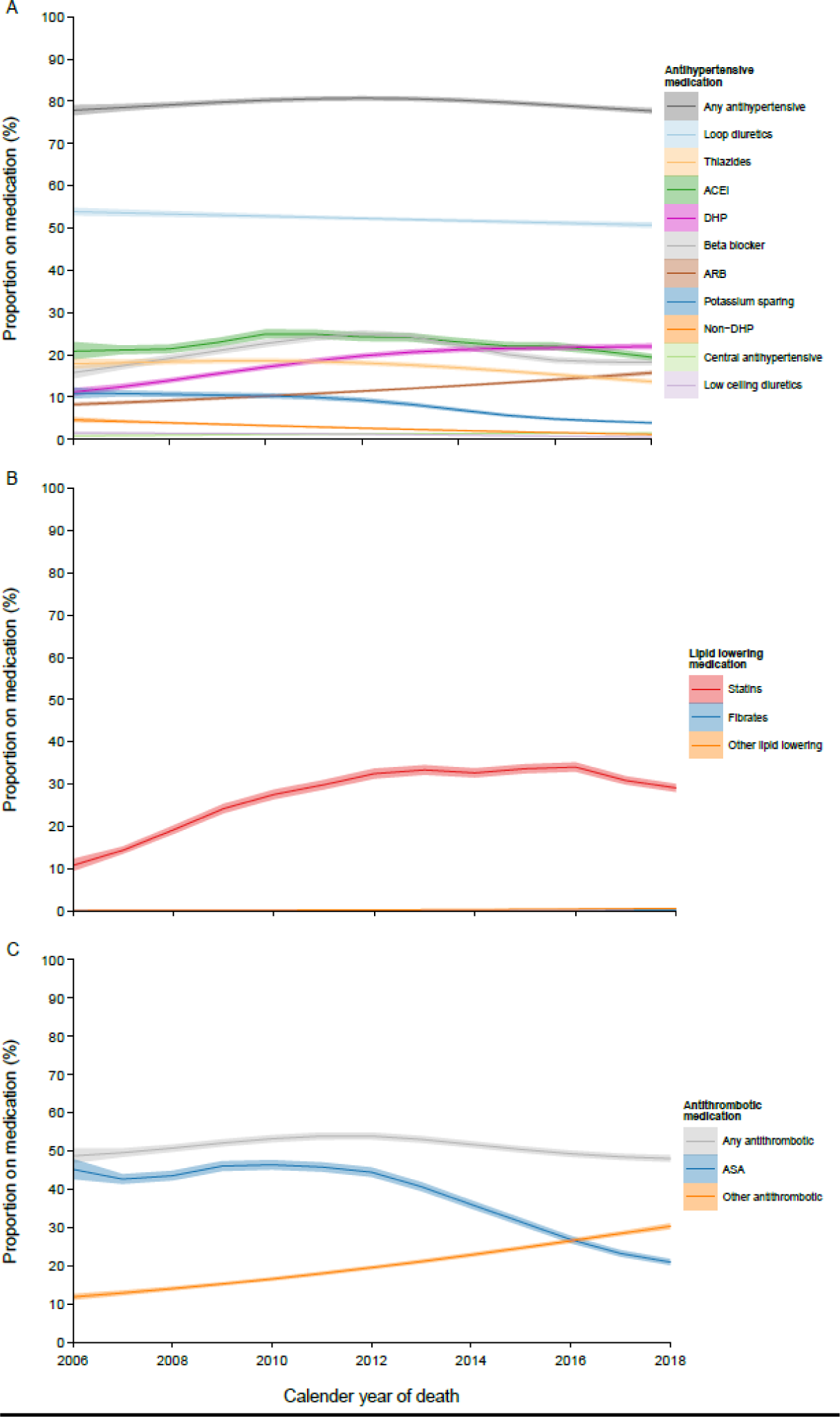
Proportion on A) antihypertensive, B) lipid lowering, and c) antithrombotic medication at time of death, by year of death and medication class. Shaded areas represent 95% CI. A crude model, without additional adjustments for covariates, was used to construct the figure. Antihypertensive medication includes loop diuretics, thiazides, angiotensin-converting enzyme inhibitors (ACEi), dihydropyridines (DHP), beta blocker, angiotensin receptor blockers (ARB), potassium-sparing diuretics, non-dihydropyridines (non-DHP), central antihypertensive, and low ceiling diuretics. Lipid lowering medication includes statins, fibrates, and other lipid lowering medication. Antithrombotic medication includes acetylsalicylic acid (ASA) and other antithrombotic medication.

Further, we analyzed covariates potentially associated with discontinuation prior to death (in the population not on medication at time of death) and being on medication at time of death by including the following variables in the Poisson and binomial models: age, diabetes duration, number of complications, level of education, country of origin, income quintile and marital status. The effect of age and diabetes duration was included in the models as a smooth parametric function. All covariates except country of origin were considered as time varying in the Poisson model. P-values less than 0.05 were considered significant. R version 4.0.2 (2020-06-22) was used for statistical analyses.

### Data approval/ethics

The study was approved by the Danish Data Protection Agency. Under Danish law, no ethic approval is required for register-based studies. All data were anonymized and located at Statistics Denmark.

## Results

A total of 52,523 (55% women) individuals with T2D were identified as deceased at age 80 or older in the period 01.01.2006 and 31.12.2018. The mean (SD) age at T2D diagnosis was 77.3 (7.8) years while the median (IQR) age and T2D duration at death was 86.5 [83.3, 90.3] and 9.2 [4.6, 14.3], respectively. Women were generally older at time of diabetes diagnosis and death and had a longer diabetes duration than men (Table 1).

**Table 1).** Characteristics at time of death for individuals with type 2 diabetes aged 80 years or older at time of death.

|  | Unit | Full sample<br>(n = 52523) | Men<br>(n = 23777 (45%)) | Women<br>(n = 28746(55%)) | p-value * |
| --- | --- | --- | --- | --- | --- |
| Age at death | Years | 86.5 [83.3, 90.3] | 85.5 [82.7, 89.0] | 87.5 [83.9, 91.3] | <0.0001 |
| Age at T2D-diagnosis | Years | 77.3 (7.8) | 76.7 (7.6) | 77.8 (7.9) | <0.0001 |
| T2D duration | Years | 9.2 [4.6, 14.3] | 8.8 [4.4, 13.8] | 9.6 [4.9, 14.7] | <0.0001 |
| No. of complications | No. (%) |  |  |  | <0.0001 |
| 0 |  | 5120 (9.7) | 1965 (8.3) | 3155 (11.0) |  |
| 1 |  | 8510 (16.2) | 3514 (14.8) | 4996 (17.4) |  |
| 2 |  | 10649 (20.3) | 4646 (19.5) | 6003 (20.9) |  |
| 3 |  | 10372 (19.7) | 4668 (19.6) | 5704 (19.8) |  |
| 4 |  | 8390 (16.0) | 3992 (16.8) | 4398 (15.3) |  |
| 5 |  | 5420 (10.3) | 2741 (11.5) | 2679 (9.3) |  |
| 6 |  | 2683 (5.1) | 1468 (6.2) | 1215 (4.2) |  |
| 7 |  | 1048 (2.0) | 590 (2.5) | 458 (1.6) |  |
| >8 |  | 331 (0.6) | 193 (0.8) | 138 (0.5) |  |
| Education | No. (%) |  |  |  | <0.0001 |
| Low |  | 26061 (49.6) | 10426 (43.8) | 15635 (54.4) |  |
| Medium |  | 11143 (21.2) | 6943 (29.2) | 4200 (14.6) |  |
| High |  | 4066 (7.7) | 2623 (11.0) | 1443 (5.0) |  |
| Missing |  | 11253 (21.4) | 3785 (15.9) | 7468 (26.0) |  |
| Country of origin † | No. (%) |  |  |  | <0.0001 |
| Danish |  | 50472 (96.1) | 22972 (96.6) | 27500 (95.7) |  |
| Non-western |  | 873 (1.7) | 401 (1.7) | 472 (1.6) |  |
| Western |  | 1178 (2.2) | 404 (1.7) | 774 (2.7) |  |
| Income quintile | No. (%) |  |  |  | <0.0001 |
| Q1 (lowest) |  | 20685 (39.4) | 8771 (36.9) | 11914 (41.5) |  |
| Q2 |  | 20849 (39.7) | 8625 (36.3) | 12224 (42.5) |  |
| Q3 |  | 5226 (10.0) | 2704 (11.4) | 2522 (8.8) |  |
| Q4 |  | 2209 (4.2) | 1361 (5.7) | 848 (3.0) |  |
| Q5 (highest) |  | 1660 (3.2) | 1094 (4.6) | 566 (2.0) |  |
| Missing |  | 1876 (3.6) | 1212 (5.1) | 664 (2.3) |  |
| Marital status | No. (%) |  |  |  | <0.0001 |
| Married |  | 14942 (28.4) | 11409 (48.0) | 3533 (12.3) |  |
| Widow/Widowed |  | 29957 (57.0) | 8827 (37.1) | 21130 (73.5) |  |
| Divorced |  | 4758 (9.1) | 2008 (8.4) | 2750 (9.6) |  |
| Single |  | 2866 (5.5) | 1533 (6.4) | 1333 (4.6) |  |
| Cardiovascular disease | No. (%) | 42247 (80.4) | 19434 (81.7) | 22813 (79.4) | <0.001 |
| Hypertensive disease | No. (%) | 30953 (58.9) | 13730 (57.7) | 17223 (59.9) | <0.001 |
| Atrial fibrillation | No. (%) | 19090 (36.3) | 8974 (37.7) | 10116 (35.2) | <0.001 |
| Heart failure | No. (%) | 16443 (31.3) | 8072 (33.9) | 8371 (29.1) | <0.001 |
| Ischemic heart disease | No. (%) | 21167 (40.3) | 10848 (45.6) | 10319 (35.9) | <0.001 |
| Macrovascular Atherosclerotic disease | No. (%) | 10075 (19.2) | 5379 (22.6) | 4696 (16.3) | <0.001 |
| Cerebrovascular disease | No. (%) | 15991 (30.4) | 7435 (31.3) | 8556 (29.8) | <0.001 |
| Diabetic kidney disease | No. (%) | 21186 (40.3) | 10362 (43.6) | 10824 (37.7) | <0.001 |
| Severe CDK | No. (%) | 5773 (11.0) | 2663 (11.2) | 3110 (10.8) | 0.169 |
| End-stage CDK | No. (%) | 2730 (5.2) | 1402 (5.9) | 1328 (4.6) | <0.001 |
| Retinopathy | No. (%) | 5540 (10.5) | 2442 (10.3) | 3098 (10.8) | 0.062 |
| Neuropathy | No. (%) | 4382 (8.3) | 2207 (9.3) | 2175 (7.6) | <0.001 |
| Amputation | No. (%) | 2910 (5.5) | 1560 (6.6) | 1350 (4.7) | <0.001 |

\*Mean (SD) was calculated for continuous variables with normal distribution, while median (Q1-Q3) was calculated for variables with right skewed distribution. Percentages were calculated for categorical variables and might not sum to 100 because of rounding. Tests for differences in means between men and women were calculated by One-way ANOVA F-test for continuous variables or with Kruskal-Wallis test when not normally distributed. Pearson's Chi-Square test of independence was used for categorical variables with expected counts above 5 in all cells.
† Western countries include EU- countries, Andorra, Australia, Canada, Iceland, Liechtenstein, Monaco, New Zealand, Norway, San Marino, Schweiz, United states of America and the Vatican. Non-western countries include Albania, Bosnia and Herzegovina, Belarus, Yugoslavia, Kosovo, Macedonia, Moldova, Montenegro, Russia, Serbia, Soviet Union, Turkey, Ukraine, and all African, Asian, Oceania countries (except Australia and New Zealand), south and central American countries and stateless.

At time of death 42,247 (80.4%) individuals had at least one CVD diagnosis, 21167 (40.3) had ischemic heart disease, 15,991 (30.4%) had cerebrovascular diseases, 21,186 (40.3%) had diabetic kidney diseases, 5,540 (10.5%) had retinopathy, 4,382 (8.3%) had neuropathy and 2,910 (5.5%) had undergone amputation. The prevalence of most complications was higher among men (Table 1).

### Extent, timing, and secular changes of discontinuation

Figure 1 shows the proportion on medication in the decade leading up to death subdivided by calendar year of death, while Figure 2 shows the proportion on medication at death as a function of year of death. Overall, the proportion on any antihypertensive medication was high and stable over time, both in the years prior to death but also with increasing calendar year of death. Discontinuations occurred predominantly during the last year of life but were initiated earlier for individuals who died in recent calendar years, although the proportion at death remained stable (78% [95% CI 77-79] vs. 78% [95% CI 77-78] for those that died in 2006 and 2018, respectively) (Figure 1A and 2A). We found that the proportion on angiotensin-converting enzyme inhibitors (ACEi) and beta-blockers increased at ten years before death, with increasing year of death. However, the proportion on both medication classes at death had an initial increase and a subsequential decrease (Figure 1A and 2A). For ACEi this reflected the relatively continuous but increased discontinuation rate at any time prior to death, whereas for beta-blockers, it was due to more pronounced discontinuation during the last year of life. Although the proportion on thiazides at ten years before death increased with calendar year of death, the earlier and increased discontinuation resulted in lower proportions at time of death for those that died in recent years (Figure 1A and 2A). Discontinuation of angiotensin II-receptor blockers (ARB) changed from primarily occurring during the last couple of years prior to death to a continuous discontinuation during the decade before death, while discontinuation of dihydropyridines (DHP) remained continuous. Discontinuation rates did not catch up with the increasing utilization of ARB and DHP, resulting in higher proportions on these drugs at any time point prior to death and at time of death, with increasing calendar year of death (Figure 1A and 2A). Furthermore, we found a decrease in the proportion on non-dihydropyridines (non-DHP), central antihypertensives and low ceiling diuretics any time prior to and at time of death, with discontinuations occurring continuously (Figure 1A and 2A). The proportion on loop-diuretics at all time points before death was high and remained stable (Figure 1A and 2A). Finally, we found a decrease in the proportion on potassium sparing medication at any point prior to and at time of death, with increasing calendar year of death (Figure 1A and 2A).

The proportion on statins markedly increased at any point prior to death with increasing calendar year of death (Figure 1B and 2B). Discontinuation trajectories changed from occurring during the last year of life to a more continuous discontinuation during the decade prior to death. This resulted in an increase in the proportion on statins at death that peaked in 2016 (from 11% [95% CI 9-12%] in 2006 to 34% [95% CI 33-35%] in 2016) followed by a subsequent decrease to 29% [95% CI 28-30%] in 2018 (Figure 2B). Similar patterns, at lower proportions, were observed for other lipid-lowering drugs (Figure S1 and S2). Both the utilization and timing of discontinuation remained unchanged for fibrates (Figure 1B and 2B).

The overall proportion on any antithrombotic medication prior to death was high and increased slightly with increasing calendar year of death. Discontinuations occurred predominantly during the last year of life, resulting in stable proportions at death (49% [95% CI 47-51%] in 2006 vs. 48% [95% CI 47-49%] in 2018) (Figure 1C and 2C). The proportion on acetylsalicylic acid (ASA) decrease while the proportion on other antithrombotic medication increased, at all times before and at time of death (Figure 1C and 2C). The timing of discontinuation of ASA changed from predominantly occurring during the last year of life to an earlier and more accelerated discontinuation. In contrast, discontinuation of other antithrombotic medication changed from a constant discontinuation during the last decade, to predominantly occurring in the last year of life (Figure 1C and 2C).

### Demographic, clinical, and socioeconomic factors associated with discontinuation

Table 2 shows the adjusted odds ratios (ORs) for selected covariates and their associations with being on medication at time of death, while table 3 shows the adjusted rate ratios (RR) of discontinuation i.e., of last medication intake, given that the date of last intake occurred prior to death. We found that women were less likely to be on statins at time of death and more likely to discontinue statins prior to death than men (Table 2 and 3). However, women were less likely to be on any antithrombotic medication, central antihypertensive medication, and ACEi at time of death and less likely to discontinue any of these medications in the decade prior to death then men. The opposite was true for most other antihypertensive medication (Table 2 and 3).

**Table 2).**
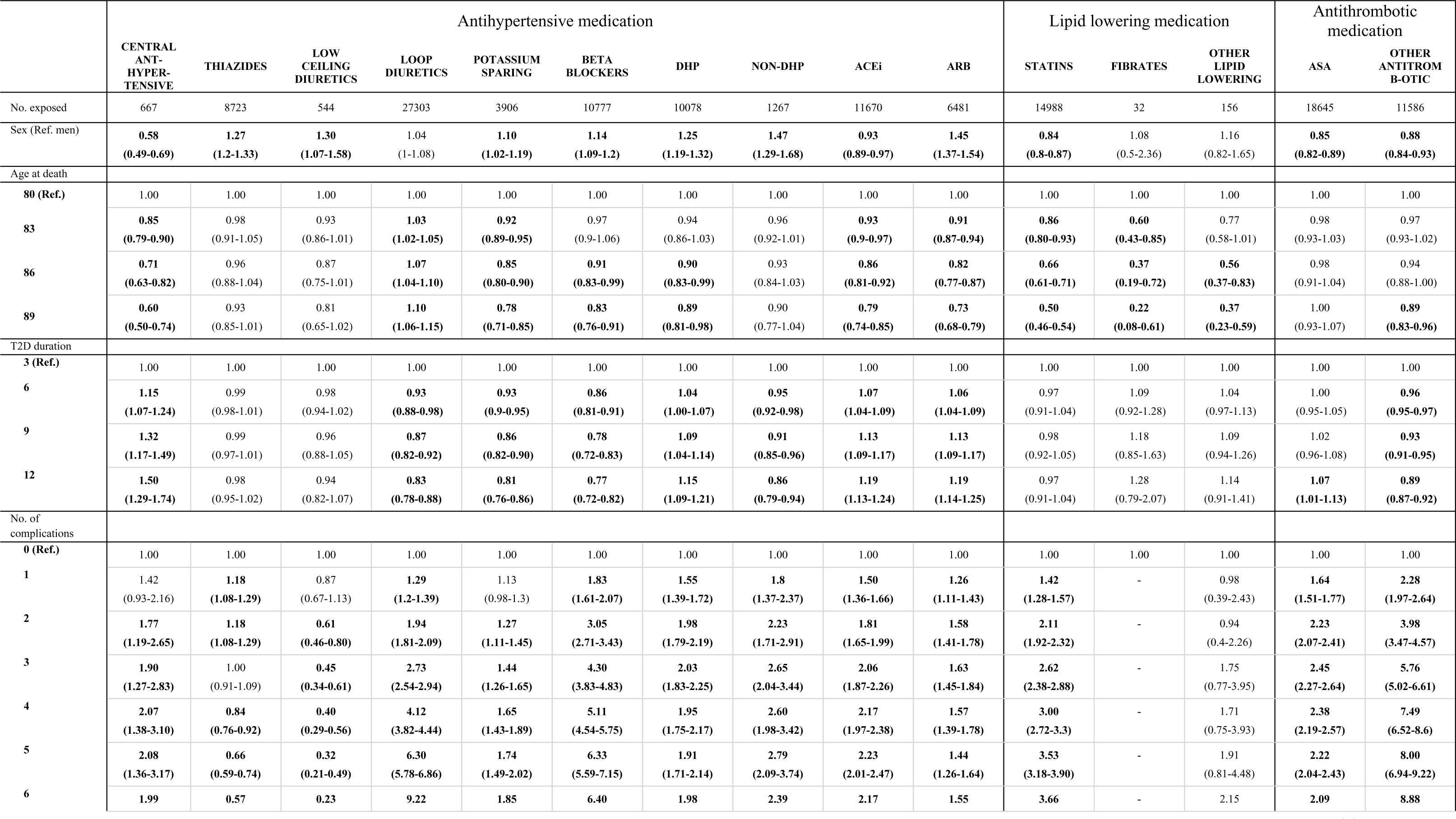
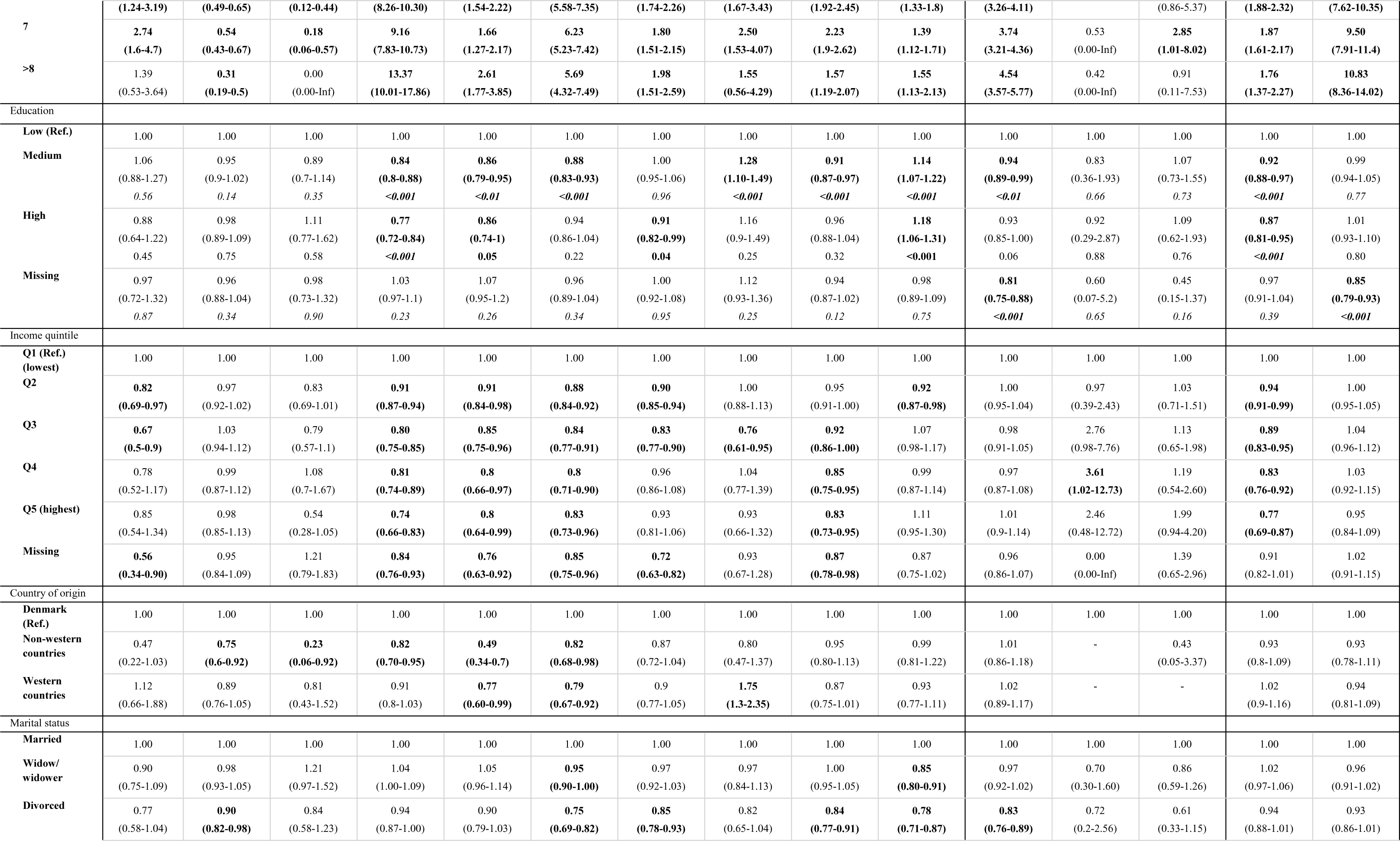
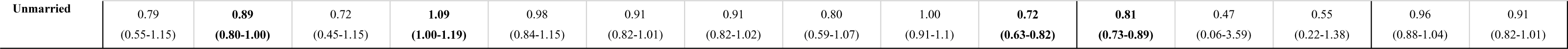
Factors associated with being <u>on</u> medication at time of death, subdivided by medication class. Results are given as adjusted odds ratios (ORs) with corresponding 95% CIs. OR in bold indicate P-values less than 0.05.

The likelihood of being on medication at time of death decreased with increasing age at death for most medication classes except for loop-diuretics, where the opposite was true. Further, we found that increasing age at death was associated with a reduced rate of discontinuing most medication classes (Table 3). Increasing T2D duration at death was associated with decreasing odds of being on loop diuretics, potassium sparing, betablockers, non-DHP, and other antithrombotic medication. Conversely, increasing T2D duration was associated with an increased odds of being on central antihypertensive medication, DHP, ACEi, and ARB. For all medication classes we found a decreased rate of discontinuation with increasing diabetes duration at death, in the decade prior to death. Overall, the presence of complications increased both the odds of being on medication at time of death and the rate of discontinuation during the ten years prior to death, except for thiazides, low ceiling diuretics, and other lipid lowering drugs than statins. Furthermore, for many of the drug classes, the odds increased with increasing number of complications.

**Table 3).**
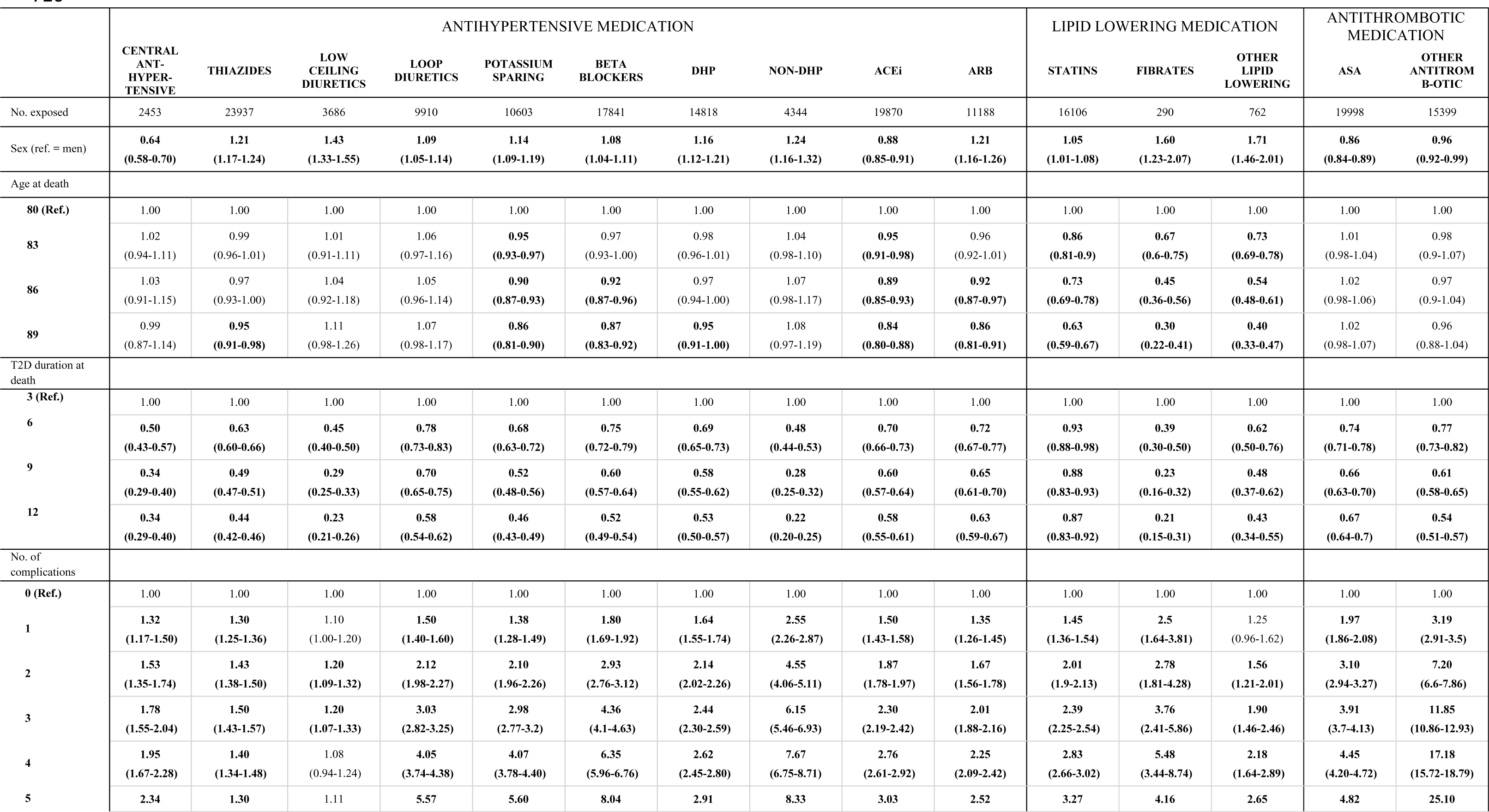
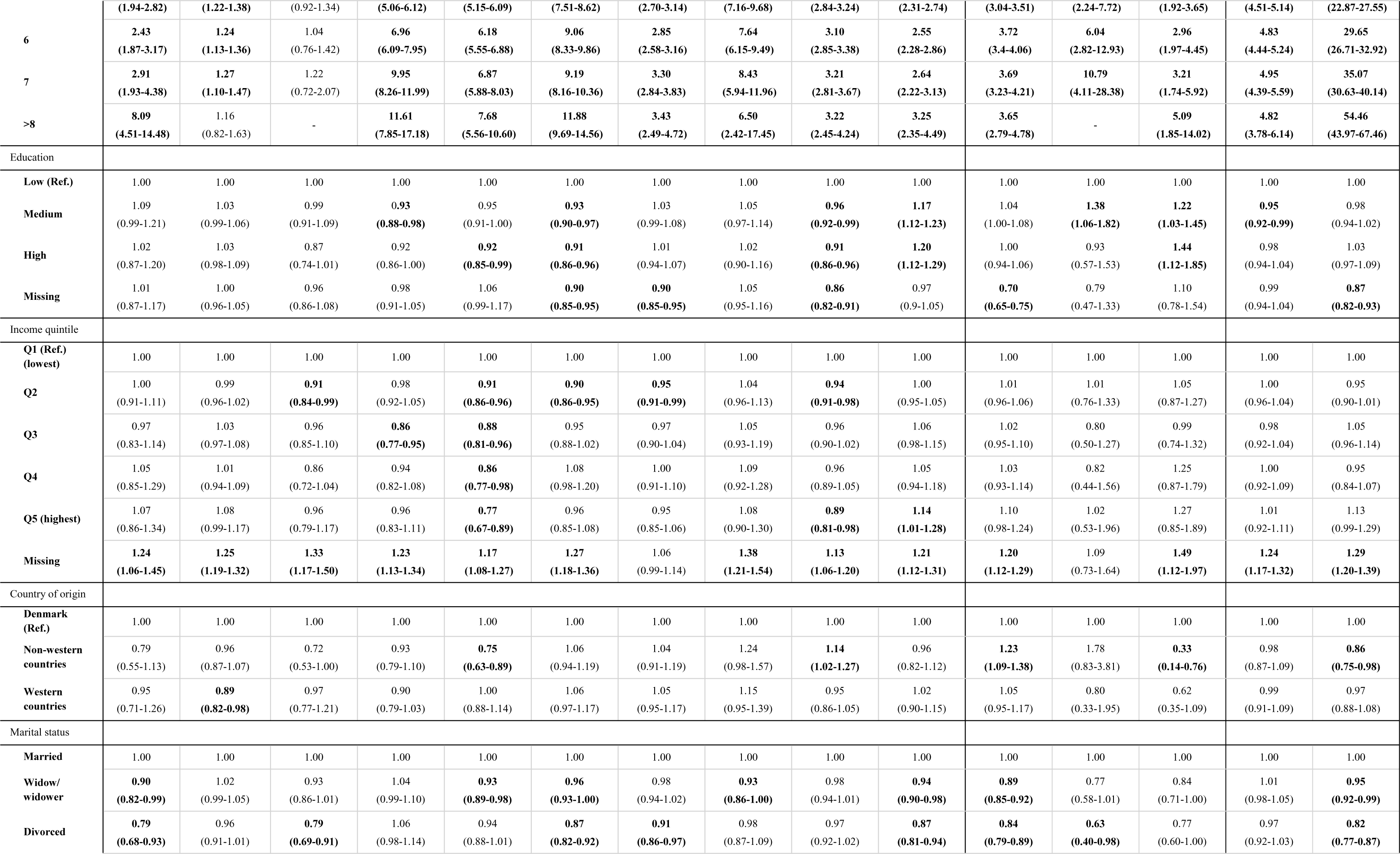
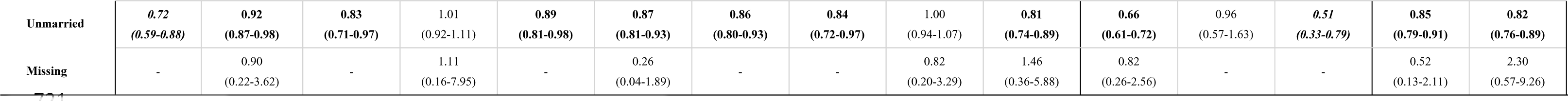
Factors associated with the rate of discontinuation in the decade prior to death, subdivided by medication class. The table contains adjusted rate ratios of discontinuation of medication i.e., intake of the last medication, assuming the date of last intake of medication occurred prior to death. Rate ratios are presented with the corresponding 95% CIs. OR in bold indicate P-values less than 0.05.

The association with sociodemographic covariates was more scattered. Higher levels of education were associated with lower odds of being on most antihypertensive medication, statins, and ASA but higher odds of being on non-DHP and ARB at time of death, compared to those with a lower level of education. Overall, a higher level of education was associated with increased discontinuation of other lipid-lowering medications and ARB and a decreased discontinuation of ACEi and beta blockers in the decade before death. Higher income quintiles were associated with decreasing odds of being on most antihypertensive medications and ASA at time of death. The association with discontinuation prior to death was less clear.

Both non-western and western immigrant status was associated with reduced odds of being on several of the anti-hypertensive medications at time of death when compared to non-immigrants. We also found that non-western immigrant status was associated with an increased rate of discontinuation of statins and ACEi in the decade before death.

Generally, being a widower, divorced or unmarried was associated with lower odds of being on statins and several of the anti-hypertensive medications, although not all the associations reached statistical significance. Finally, widowers, divorced and unmarried individuals were less likely to discontinue other antithrombotic medication, statins, central antihypertensive medication, beta-blockers, and ARB compared to married individuals in the ten years prior to death.

## Discussion

Based on Danish nationwide registers, we have described the extent, timing, and secular changes of discontinuation of cardioprotective medication in a deceased population of elderly with T2D, during the last decade of life. Furthermore, we have explored the association of various covariates with these patterns prior to and at time of death.

The increased focus on reducing CVD morbidity and mortality in the elderly population with T2D has prompted an increased use of cardioprotective medications for both primary and secondary prevention. This is reflected in our results by the increasing proportion receiving these medications. Previous studies on prescription trends of cardioprotective drugs in the very elderly (>80 years) with T2D have yielded similar trends.^24,25^

With growing evidence for the effectiveness of hypertension treatment in reducing CVD morbidity and mortality in older adults, the use of ACEi, ARB, beta-blockers, thiazides and DHP has intensified.^26^ ARB and ACEi are considered as first-line therapy in T2D and are interchangeable when one is not tolerated.^5,27^ Historically in Denmark, ARB’s have been favored over ACEi due to insufficient evidence supporting the latter in T2D in the early 2000s.^28^ However, higher proportions on ACE compared to ARB might be explained by time-trends in recommendation, marketing, and price. ACEís were introduced 15 years before ARB and have consequently been both cheaper and more well-documented (in a non-diabetes population) during the start of the study period. In later years, with equalizing prices and compiling evidence for ARB there has been an increased utilization of ARB and reflexive discontinuation of ACEi. Further, the discontinuation rates may also be influenced by declining kidney function and/or blood pressure and adherence.^29^

Increasing age and diabetes duration at death were negatively associated with the proportions on ACEi, ARB, betablockers and potassium sparing drugs, however, increasing number of complications were positively associated with the proportion on all antihypertensive medications. These associations can partly be explained by a combination of a reduced absolute risk reduction of the benefits of treatment in those over 80 years, the increased focus on individual treatment goals (considering both chronological and biological age) and the elevated risk of side effects in this population.^26,30,31^ Common side effects to antihypertensive medication include cough, dizziness, flushing, palpitations, dehydration, and electrolyte disturbances, all known facilitators of discontinuation.^32,33^ Unfortunately, in this study we had no information on the degree of side effects or the reason of discontinuation. Moreover, indications for antihypertensive medication include a broad-spectrum of prevalent diseases in this population, such as heart failure, arrythmias, angina pectoris and myocardial infarctions, all of which are likely to contribute to the increased utilization while not being appropriate for discontinuation.^34^

Use of lipid-lowering medication in the elderly is still a subject for discussion, nevertheless, we found a considerable increase in statin use, in line with previous studies.^24,25,35^ Furthermore, we found that increasing number of complications was positively associated with the proportion on statins in the decade prior to death (expressed as an increased rate of discontinuation), suggesting that use of statins may have primarily served as secondary prevention. This is in accordance with evidence suggesting that lipid-lowering drugs (statins in particular) are effective in reducing CVD in elderly (>75 years) when used as secondary prevention, however, the evidence for primary prevention is less clear.^36,37^ The increased and earlier discontinuation of statins found in the present study is deemed to be multifactorial and driven by non-adherence (especially in primary prevention), increasing age and the negative attention drawn to potential side effects, both serious and not.^38^ Evidence on this field proposes a low risk for serious side effects and rejects that milder symptoms such as muscle pains are related to statins.^39–41^

The decline in the proportion on ASA with increasing calendar year of death could partly be attributed to the uncertainty of the absolute net value of ASA as primary prevention and the subsequent shift in recommendation, from T2D being a singular risk-factor to inclusion of high CVD risk for primary prevention.^25,42^ Another factor might be the increased attention on the risk of gastrointestinal bleeding, which is particularly pronounced in the elderly population.^43^ Further, the positive association with increasing number of complications seen for antithrombotic medication indicates use as secondary prevention.

In this study, we did not distinguish between other types of antithrombotic medication, thus, we cannot determine which medication class/classes are responsible for driving the increased use. Results from a study, including one region in Denmark, found an increase in direct oral anticoagulants and clopidogrel, and a decrease in vitamin-k antagonist and other antiplatelets.^25^ The time-limited nature of some of these medications, such as clopidogrel and ticagrelor after coronary stent implantations, may further contribute to the discontinuation rates.^44^

In Denmark, health care equality is promoted by free access to health care and gradual reimbursements of medication, where the extent of reimbursement increases with the quantity of medication purchased. Still, medication prices can influence the choice of medication, but we found no evidence of that in our study.

### End of life

When approaching end of life, it is recommended to change treatment strategies towards symptom relief and comfort. Specific discontinuation guidelines are also provided by the Danish health authority’s recommending discontinuation of statins with limited life expectancy (independent of prevention type). Similarly, discontinuation of ASA is recommended in the absence of CVD, avoidance of loop-diuretics in uncomplicated hypertension unless accompanied by peripheral edema, and consideration of discontinuing beta blockers two years post myocardial infarction.^45^ The remaining medication classes lack specific recommendations. Our finding of pronounced discontinuations during the last years of life and more so in the recent years, may indicate that these recommendations are increasingly adhered to.^5^ Discontinuations may have been additionally prompted by decreasing blood pressure, cholesterol levels or kidney function, dependent or independent of the anticipation of death.^29,46^ We found that the likelihood of being on medication at time of death increased with the presence of complications for all medications and diabetes duration for some of the antihypertensive medications. In line with our results, Engell et al, found an increased likelihood of discontinuation with advancing age but a reduced likelihood with the presence of comorbidities.^47^

The proportion on antihypertensive and antithrombotic medication at death remained stable throughout the study period, while the proportion on statins increased, though with a slight reduction in current years. This resulted in 77% [95% CI 77-78%] of individuals on any antihypertensive medication, 29% [95% CI 28-30%] on statins and 48% [95% CI 47-49%] on any antithrombotic medication at time of death in 2018. The targeted proportion on these medication at time of death is unknown, however, the appropriateness of medication is cardinal.^48^ Unfortunately, our study cannot provide insights into the appropriateness of medication. In accordance with our study, though with differing proportions, Hamada et al found a decrease in proportion on antihypertensives from 76% to 68%, statins from 62% to 54%, and low-dose aspirin from 46% to 42%, between the fourth quarter and the first quarter prior to death.^49^

There are several barriers to discontinuation of cardioprotective medication that may influence our results, concerning both patients and health care professionals. Common barriers among health care professionals include uncertainty of the risk-benefit balance of the medication, lack of evidence of the effect of discontinuation, uncertainty of timing of discontinuation and limited training. Patient reported barriers include lack of understanding of the risk-benefit of medication, risk and risk reduction of CVD, and the ability to actively control the progression of their disease.^11^ Therefore, an unavoidable conclusion is the unmet need for a greater inclusion of elderly individuals in RCTs on the efficiency and safety of medication, and a general need for RCTs testing timing and safety of discontinuation.

### Strength and limitations

There are several strengths of this study. A major strength is the use of reliable data derived from valid and complete nationwide registers. In particular, the use of the DMreg enabled us to gather a representative cohort of elderly with a T2D diagnosis treated in both outpatient clinics and general practice (GP), with minimal selection bias. This was of particular importance as approximately 80% of the T2D population is treated in GP and therefore not necessarily be registered in the DNPR. Another strength was the use of the NPR, which enabled the identification of all redeemed prescriptions and thereby eliminating any recall bias.

There are some potential limitations to our study. Although we adjusted models for several covariates with possible influence on timing of last intake of medication, there are still several covariates that we were unable to access. These include frailty indicators, self-management abilities, laboratory measurements (electrolytes, cholesterol, kidney, and liver function), reports on adverse effects, and clinical measurements (blood pressure). Another possible limitation is the absence of hospital dispensed medication. This is a potential source of misclassification of the date of discontinuation (both under- and overestimation) as there is an increased prevalence of hospitalization in this population when approaching death, resulting in a substantial proportion dying in the hospital. The timing of discontinuation is based on several assumptions, one being that redeemed medication is consumed as prescribed. Unfortunately, the degree of missingness of the prescribed dose was substantial and thus the Defined Daily Dose (as defined by the WHO) had to be used as a proxy, adding to the risk of misclassification. Finally, as hypertensive disease and mild forms of retinopathy and neuropathy are underreported in in the DNPR, and macrovascular arteriosclerotic disease is underdiagnosed. Thus, the associations related to the comorbidity variable may be underestimated.

## Conclusion

In conclusion, we found an increase in the proportion on cardioprotective medication among elderly with T2D from 2006 to 2018, which was primarily driven by an increased use of ACEi, ARB, DHP, beta blockers and statins. Discontinuations shifted towards earlier discontinuation but occurred continuously and were particularly pronounced during the last year of life. Sex, age, diabetes duration and number of complications were associated with the proportion on medication in the decade prior to and at time of death, however, the direction of the associations varied between medication classes. Sociodemographic covariates had a more scattered association with the outcome. Future studies on timing and safety of discontinuation, as well as inclusion of a more representative elderly population in RCTs assessing the efficiency and safety of medication is needed for closing the current knowledge gaps in this field.

## Contribution statement

VK, BC, GSA, JR, BB and HRC contributed to the conception of the study. BC detailed the statistical method and VK performed the analyses, was responsible for data-management and contributed to obtaining data. VK wrote the initial draft of the manuscript and was responsible for visualization of results. GSA, JR, BC, MEJ, BB, HRC and SHS contributed to revising the article and supervision. VK, BC, SHS and JR contributed with interpretation of data. VK is the guarantor of this work and, as such, had full access to all the data in the study and takes responsibility for the integrity of the data and the accuracy of the data analysis. The authors declare that the results of the study are presented clearly, honestly, and without fabrication, falsification, or inappropriate data manipulation.

## Data Availability

Study data cannot and therefore will not be made publicly available as all data is held at Statistics Denmark's servers, and individual data from these are confidential for data privacy reasons. Access to data requires an application and permissions to access the patient level data.

## Acknowledgements

The authors thank the Danish Clinical Registries (RKKP) for giving permission to use clinical data for this study.

## Supplementary material

Expanded Methods

Tables S1-S2

Figure S1-S2

## References

1. World Health O. World report on ageing and health. Geneva: World Health Organization; 2015.

2. Carstensen B, Rønn PF, Jørgensen ME. Prevalence, incidence and mortality of type 1 and type 2 diabetes in Denmark 1996-2016. BMJ open diabetes research & care. 2020;8. doi: 10.1136/bmjdrc-2019-001071

3. Rosella LC, Negatu E, Kornas K, Chu C, Zhou L, Buajitti E. Multimorbidity at time of death among persons with type 2 diabetes: a population-based study in Ontario, Canada. BMC Endocr Disord. 2023;23:127. doi: 10.1186/s12902-023-01362-x

4. Huang ES, Laiteerapong N, Liu JY, John PM, Moffet HH, Karter AJ. Rates of complications and mortality in older patients with diabetes mellitus: the diabetes and aging study. JAMA internal medicine. 2014;174:251–258. doi: 10.1001/jamainternmed.2013.12956

5. ElSayed NA, Aleppo G, Aroda VR, Bannuru RR, Brown FM, Bruemmer D, Collins BS, Hilliard ME, Isaacs D, Johnson EL, et al. 13. Older Adults: Standards of Care in Diabetes— 2023. Diabetes care. 2022;46:S216–S229. doi: 10.2337/dc23-S013

6. Rossello X, Pocock SJ, Julian DG. Long-Term Use of Cardiovascular Drugs: Challenges for Research and for Patient Care. J Am Coll Cardiol. 2015;66:1273–1285. doi: 10.1016/j.jacc.2015.07.018

7. Wastesson JW, Morin L, Tan ECK, Johnell K. An update on the clinical consequences of polypharmacy in older adults: a narrative review. Expert Opin Drug Saf. 2018;17:1185–1196. doi: 10.1080/14740338.2018.1546841

8. Remelli F, Ceresini MG, Trevisan C, Noale M, Volpato S. Prevalence and impact of polypharmacy in older patients with type 2 diabetes. Aging Clin Exp Res. 2022;34:1969–1983. doi: 10.1007/s40520-022-02165-1

9. Wu H, Kouladjian O’Donnell L, Fujita K, Masnoon N, Hilmer SN. Deprescribing in the Older Patient: A Narrative Review of Challenges and Solutions. Int J Gen Med. 2021;14:3793–3807. doi: 10.2147/ijgm.S253177

10. Johansson KS, Kornholt J, Bülow C, Petersen TS, Perrild H, Rungby J, Christensen MB. Physician-led medication reviews in polypharmacy patients treated with at least 12 medications in a type 2 diabetes outpatient clinic: A randomised trial. Diabetic medicine : a journal of the British Diabetic Association. 2023;40:e15052. doi: 10.1111/dme.15052

11. Brunner L, Rodondi N, Aubert CE. Barriers and facilitators to deprescribing of cardiovascular medications: a systematic review. BMJ open. 2022;12:e061686. doi: 10.1136/bmjopen-2022-061686

12. Hickman E, Seawoodharry M, Gillies C, Khunti K, Seidu S. Deprescribing in cardiometabolic conditions in older patients: a systematic review. Geroscience. 2023. doi: 10.1007/s11357-023-00852-z

13. Sinclair A, Dunning T., Colagiuri, S. IDF Global Guideline For Manging Older People With Type 2 Diabetes. International Diabetes Federation. https://www.idf.org/e-library/guidelines/78-global-guideline-for-managing-older-people-with-type-2-diabetes.html. 2013.

14. Oktora MP, Kerr KP, Hak E, Denig P. Rates, determinants and success of implementing deprescribing in people with type 2 diabetes: A scoping review. Diabetic medicine : a journal of the British Diabetic Association. 2021;38:e14408. doi: 10.1111/dme.14408

15. Schmidt M, Pedersen L, Sorensen HT. The Danish Civil Registration System as a tool in epidemiology. European journal of epidemiology. 2014;29:541–549. doi: 10.1007/s10654-014-9930-3

16. Pottegård A, Schmidt SAJ, Wallach-Kildemoes H, Sørensen HT, Hallas J, Schmidt M. Data Resource Profile: The Danish National Prescription Registry. Int J Epidemiol. 2017;46:798–798f. doi: 10.1093/ije/dyw213

17. Schmidt M, Schmidt SA, Sandegaard JL, Ehrenstein V, Pedersen L, Sørensen HT. The Danish National Patient Registry: a review of content, data quality, and research potential. Clin Epidemiol. 2015;7:449–490. doi: 10.2147/clep.S91125

18. Sundhedsdatastyrelsen. Laboratoriedatabasen. https://sundhedsdatastyrelsen.dk/da/registre-og-services/om-de-nationale-sundhedsregistre/doedsaarsager-og-biologisk-materiale/laboratoriedatabasen. 01.09.2020. Accessed 19–10.

19. Jorgensen ME, Kristensen JK, Reventlov Husted G, Cerqueira C, Rossing P. The Danish Adult Diabetes Registry. Clin Epidemiol. 2016;8:429–434. doi: 10.2147/clep.s99518

20. Jensen VM, Rasmussen AW. Danish education registers. Scandinavian journal of public health. 2011;39:91–94. doi: 10.1177/1403494810394715

21. Pedersen CB. The Danish Civil Registration System. Scandinavian journal of public health. 2011;39:22–25. doi: 10.1177/1403494810387965

22. Baadsgaard M, Quitzau J. Danish registers on personal income and transfer payments. Scandinavian journal of public health. 2011;39:103–105. doi: 10.1177/1403494811405098

23. Zanders MM, van Steenbergen LN, Haak HR, Rutten HJ, Pruijt JF, Poortmans PM, Lemmens VE, van de Poll-Franse LV. Diminishing differences in treatment between patients with colorectal cancer with and without diabetes: a population-based study. Diabetic medicine : a journal of the British Diabetic Association. 2013;30:1181–1188. doi: 10.1111/dme.12253

24. Hamada S, Gulliford MC. Antidiabetic and cardiovascular drug utilisation in patients diagnosed with type 2 diabetes mellitus over the age of 80 years: a population-based cohort study. Age Ageing. 2015;44:566–573. doi: 10.1093/ageing/afv065

25. García Rodríguez LA, Cea Soriano L, de Abajo FJ, Valent F, Hallas J, Gil M, Cattaruzzi C, Rodriguez-Martin S, Vora P, Soriano-Gabarró M, et al. Trends in the use of oral anticoagulants, antiplatelets and statins in four European countries: a population-based study. European journal of clinical pharmacology. 2022;78:497–504. doi: 10.1007/s00228-021-03250-6

26. Musini VM, Tejani AM, Bassett K, Puil L, Wright JM. Pharmacotherapy for hypertension in adults 60 years or older. The Cochrane database of systematic reviews. 2019;6:Cd000028. doi: 10.1002/14651858.CD000028.pub3

27. Hansen K.B. KJK, Balasubramaniam K., Bjerregaard-Andersen M., Breum, M. Charles L., Hansen L.J., Kielgast U. KV, Madsen G.K., Bruun J. M., Navntoft D., Pietraszek A., Rossing P., Rungby J., Jensen M. S.,, Snorgaard O. SJV, Søndergaard E., Knudsen S. T. Type 2 Diabetes. https://endocrinology.dk/nbv/diabetes-melitus/behandling-og-kontrol-af-type-2-diabetes/. 2023. Accessed 06.09.2023.

28. Catalá-López F, Macías Saint-Gerons D, González-Bermejo D, Rosano GM, Davis BR, Ridao M, Zaragoza A, Montero-Corominas D, Tobías A, de la Fuente-Honrubia C, et al. Cardiovascular and Renal Outcomes of Renin-Angiotensin System Blockade in Adult Patients with Diabetes Mellitus: A Systematic Review with Network Meta-Analyses. PLoS medicine. 2016;13:e1001971. doi: 10.1371/journal.pmed.1001971

29. Ravindrarajah R, Hazra NC, Hamada S, Charlton J, Jackson SHD, Dregan A, Gulliford MC. Systolic Blood Pressure Trajectory, Frailty, and All-Cause Mortality >80 Years of Age: Cohort Study Using Electronic Health Records. Circulation. 2017;135:2357–2368. doi: 10.1161/circulationaha.116.026687

30. Bejan-Angoulvant T, Saadatian-Elahi M, Wright JM, Schron EB, Lindholm LH, Fagard R, Staessen JA, Gueyffier F. Treatment of hypertension in patients 80 years and older: the lower the better? A meta-analysis of randomized controlled trials. J Hypertens. 2010;28:1366–1372. doi: 10.1097/HJH.0b013e328339f9c5

31. de Boer IH, Bangalore S, Benetos A, Davis AM, Michos ED, Muntner P, Rossing P, Zoungas S, Bakris G. Diabetes and Hypertension: A Position Statement by the American Diabetes Association. Diabetes care. 2017;40:1273–1284. doi: 10.2337/dci17-0026

32. Conroy SP, Westendorp RGJ, Witham MD. Hypertension treatment for older people— navigating between Scylla and Charybdis. Age and Ageing. 2018;47:505–508. doi: 10.1093/ageing/afy053

33. Sommerauer C, Kaushik N, Woodham A, Renom-Guiteras A, Martinez YV, Reeves D, Kunnamo I, Al Qur An T, Hübner S, Sönnichsen A. Thiazides in the management of hypertension in older adults - a systematic review. BMC Geriatr. 2017;17:228. doi: 10.1186/s12877-017-0576-3

34. K. Brøsen US, J.P. Kampmann, S. Thirstrup Basal og klinisk farmakologi. FADL’s Forlag; 2014.

35. Thompson W, Jarbøl DE, Nielsen JB, Haastrup P, Pottegård A. Statin use and discontinuation in Danes age 70 and older: a nationwide drug utilisation study. Age Ageing. 2021;50:554–558. doi: 10.1093/ageing/afaa160

36. Gencer B, Marston NA, Im K, Cannon CP, Sever P, Keech A, Braunwald E, Giugliano RP, Sabatine MS. Efficacy and safety of lowering LDL cholesterol in older patients: a systematic review and meta-analysis of randomised controlled trials. *Lancet (London*, England). 2020;396:1637–1643. doi: 10.1016/s0140-6736(20)32332-1

37. Efficacy and safety of statin therapy in older people: a meta-analysis of individual participant data from 28 randomised controlled trials. Lancet (London, England). 2019;393:407–415. doi: 10.1016/s0140-6736(18)31942-1

38. Kriegbaum M, Liisberg KB, Wallach-Kildemoes H. Pattern of statin use changes following media coverage of its side effects. Patient Prefer Adherence. 2017;11:1151–1157. doi: 10.2147/ppa.S133168

39. Nanna MG, Abdullah A, Mortensen MB, Navar AM. Primary prevention statin therapy in older adults. Curr Opin Cardiol. 2023;38:11–20. doi: 10.1097/hco.0000000000001003

40. Thompson W, Jarbøl DE, Haastrup P, Nielsen JB, Pottegård A. Statins in Older Danes: Factors Associated With Discontinuation Over the First 4 Years of Use. Journal of the American Geriatrics Society. 2019;67:2050–2057. doi: 10.1111/jgs.16073

41. Gulliford M, Ravindrarajah R, Hamada S, Jackson S, Charlton J. Inception and deprescribing of statins in people aged over 80 years: cohort study. Age Ageing. 2017;46:1001–1005. doi: 10.1093/ageing/afx100

42. Baigent C, Blackwell L, Collins R, Emberson J, Godwin J, Peto R, Buring J, Hennekens C, Kearney P, Meade T, et al. Aspirin in the primary and secondary prevention of vascular disease: collaborative meta-analysis of individual participant data from randomised trials. *Lancet (London*, England). 2009;373:1849–1860. doi: 10.1016/s0140-6736(09)60503-1

43. Rosberg V, Grove EL, Bøtker HE, Kristensen SD, Pareek M. [Acetylsalicylic acid for primary prevention of cardiovascular events in patients with Type 2 diabetes mellitus]. Ugeskrift for laeger. 2019;181.

44. Krishnaswami A, Steinman MA, Goyal P, Zullo AR, Anderson TS, Birtcher KK, Goodlin SJ, Maurer MS, Alexander KP, Rich MW, et al. Deprescribing in Older Adults With Cardiovascular Disease. Journal of the American College of Cardiology. 2019;73:2584–2595. doi: doi:10.1016/j.jacc.2019.03.467

45. autority Dh. Seponeringslisten 2023. https://www.sst.dk/da/viden/Laegemidler/Rationel-Farmakoterapi/Medicingennemgang/Seponeringslisten. Accessed 8 SEP 2023.

46. Charlton J, Ravindrarajah R, Hamada S, Jackson SH, Gulliford MC. Trajectory of Total Cholesterol in the Last Years of Life Over Age 80 Years: Cohort Study of 99,758 Participants. *The journals of gerontology Series A*, Biological sciences and medical sciences. 2018;73:1083–1089. doi: 10.1093/gerona/glx184

47. Engell AE, Bathum L, Andersen JS, Thompson W, Lind BS, Jørgensen HL, Nexøe J. Factors associated with statin discontinuation near end of life in a Danish primary health care cohort. Fam Pract. 2023;40:300–307. doi: 10.1093/fampra/cmac090

48. Halliday BP, Wassall R, Lota AS, Khalique Z, Gregson J, Newsome S, Jackson R, Rahneva T, Wage R, Smith G, et al. Withdrawal of pharmacological treatment for heart failure in patients with recovered dilated cardiomyopathy (TRED-HF): an open-label, pilot, randomised trial. *Lancet (London*, England). 2019;393:61–73. doi: 10.1016/s0140-6736(18)32484-x

49. Hamada S, Gulliford MC. Drug prescribing during the last year of life in very old people with diabetes. Age Ageing. 2017;46:147–151. doi: 10.1093/ageing/afw174

## Supplementary references

1. Carstensen B, Rønn PF, Jørgensen ME. Prevalence, incidence and mortality of type 1 and type 2 diabetes in Denmark 1996-2016. BMJ open diabetes research & care. 2020;8. doi: 10.1136/bmjdrc-2019-001071

2. Schmidt M, Pedersen L, Sorensen HT. The Danish Civil Registration System as a tool in epidemiology. European journal of epidemiology. 2014;29:541–549. doi: 10.1007/s10654-014-9930-3

3. Pottegård A, Schmidt SAJ, Wallach-Kildemoes H, Sørensen HT, Hallas J, Schmidt M. Data Resource Profile: The Danish National Prescription Registry. Int J Epidemiol. 2017;46:798–798f. doi: 10.1093/ije/dyw213

4. Schmidt M, Schmidt SA, Sandegaard JL, Ehrenstein V, Pedersen L, Sørensen HT. The Danish National Patient Registry: a review of content, data quality, and research potential. Clin Epidemiol. 2015;7:449–490. doi: 10.2147/clep.S91125

5. Andersen JS, Olivarius Nde F, Krasnik A. The Danish National Health Service Register. Scandinavian journal of public health. 2011;39:34–37. doi: 10.1177/1403494810394718

6. Jørgensen ME, Kristensen JK, Reventlov Husted G, Cerqueira C, Rossing P. The Danish Adult Diabetes Registry. Clin Epidemiol. 2016;8:429–434. doi: 10.2147/clep.s99518

7. Andersen N, Hjortdal J, Schielke KC, Bek T, Grauslund J, Laugesen CS, Lund-Andersen H, Cerqueira C, Andresen J. The Danish Registry of Diabetic Retinopathy. Clin Epidemiol. 2016;8:613–619. doi: 10.2147/clep.s99507

8. Baadsgaard M, Quitzau J. Danish registers on personal income and transfer payments. Scandinavian journal of public health. 2011;39:103–105. doi: 10.1177/1403494811405098

